# Causal Debiasing for Unknown Bias in Histopathology - A Colon Cancer Use Case

**DOI:** 10.1101/2024.04.25.24306407

**Authors:** Ramón L. Correa-Medero, Rish Pai, Kingsley Ebare, Daniel D. Buchanan, MarkA. Jenkins, Amanda I. Phipps, Polly A. Newcomb, Steven Gallinger, Robert Grant, Loic Le marchand, Imon Banerjee

## Abstract

Advancement of AI has opened new possibility for accurate diagnosis and prognosis using digital histopathology slides which not only saves hours of expert effort but also makes the estimation more standardized and accurate. However, preserving the AI model performance on the external sites is an extremely challenging problem in the histopathology domain which is primarily due to the difference in data acquisition and/or sampling bias. Although, AI models can also learn spurious correlation, they provide unequal performance across validation population. While it is crucial to detect and remove the bias from the AI model before the clinical application, the cause of the bias is often *unknown*. We proposed a *Causal Survival model* that can reduce the effect of unknown bias by leveraging the causal reasoning framework. We use the model to predict recurrence-free survival for the colorectal cancer patients using quantitative histopathology features from seven geographically distributed sites and achieve equalized performance compared to the baseline traditional Cox Proportional Hazards and DeepSurvival model. Through ablation study, we demonstrated benefit of novel addition of latent probability adjustment and auxiliary losses. Although detection of cause of unknown bias is unsolved, we proposed a causal debiasing solution to reduce the bias and improve the AI model generalizibility on the histopathology domain across sites.

## 1. Introduction

Histopathology analysis is usually needed when a definitive diagnosis is required, and digitization of standard tissue glass slides into Whole Slide Images (WSIs) has opened a new possibility for the use of advanced computer vision techniques not only for diagnosis but also for prognosis of advanced diseases [1, 2]. Given the complexity of histologic image interpretation and uncertain prognosis on typical histologic imaging appearance [3], colorectal cancer can be an ultimate use case for computer vision not only for diagnosis but also for prognosis, where an artificial intelligence (AI) model will be trained on a large sample of data to learn pattern indicative of poor prognosis. The primary goal of training an AI model is to make the process accurate and less time consuming for human pathologists, and the model can be applied to a wider population where the availability of expert pathologist may be limited [2].

Evaluation of model generalization play a critical role in current AI system which is usually performed by validating the model performance on an external dataset [4]. However, the accuracy of the model is often significantly degraded on the external dataset as reported by many studies in literature [5]. Performance drops are often found to be caused by models being biased to spurious patterns in the training data [5]. As a result, many machine learning methods are unsuccessful on unseen data from similar domains while they achieve highly promising results on training or test sets within the same domain. In addition, the decisions of an AI algorithm reflect discriminatory behavior toward certain majority groups or populations as they are heavily represented in the training group [6].

In a recent study, Howard et al. [7] reported that some models trained on TCGA datasets detect source sites instead of predicting prognosis or mutation states that is neither expected nor desired. Such performance could be based on the fact that the distribution of clinical information, such as survival and gene expression patterns, significantly differs among the samples provided by various laboratories. Many authors [8] considered differences in slide staining as a primary factor for the imaging bias and tried to solve it by color augmentation and stain-normalization [9, 10, 11]. In addition to acquisition specific bias, such as scanner configuration and noise, stain variation, and artifacts, bias may exist in the targeted population. For example, Zech et.al. [12] showed that AI can diagnose pneumonia from radiology images, but it uses portable intensive care unit radiographic markers as surrogates. Similarly, Rueckel et al. found a pneumothorax detection model that used shortcuts based on inserted chest tubes [13]. It has also been observed that imaging AI models learn spurious age, sex, and race correlations from images when trained for seemingly unrelated tasks [14]. More concerning direction, studies have shown AI imaging models demonstrate bias against historically underserved subgroups of age, sex, and race in disease diagnosis [15].

Given the complexity of data collection and risk of associate bias, a significant amount of work has been done to solve bias using computational pipeline [16]. Although, most of the models are either trained with the adversarial branch or applied a privacy preserving approach for reducing bias, it is only applicable when the cause of the bias is known and is a collectable variable. The bias detection and recognition is particularly challenging when faced with multiple types of unknown biases.

Alabdoulmohsin et al. [17] tackled the challenge of data sampling bias as a domain adaptation problem. The authors argue a sub-population defined by a hidden variable *U* has varying prevalence across institutions. Such a population also has varying relations with disease occurrence *Y*, and the image features *X* due to different social determinants of health that influence outcomes through comorbidities and access to care itself. They proposed an unsupervised methodology to estimate the latent population in new datasets and, therefore, adjust model predictions, lessening the effect of the shift on the model performance.

In this work, we propose a novel survival model by incorporating the concept of the latent-shift method to reduce the effect of unknown bias by treating the generalization as a domain adaptation problem. We followed Alabdulmohsin et al. [17] unsupervised domain adaptation technique to the latent distribution shifts, which generalize the standard settings of covariate aed label shift of the domain. We leverage auxiliary data in the source domain as concepts and a proxy, and generalizing it to derive an identification strategy for the optimal predictor under the target distribution. We extend the domain adaptation strategy for deep latent feature space using the auxiliary variables - cancer stage and proxy for socioeconomic status - treatment sites.

## 2. Methods

### 2.1. Dataset

The study population consisted of patients with colorectal carcinoma from the Colon Cancer Family Registry (CCFR, participating sites: Seattle, Australia, Mayo Clinic, Ontario, and Hawaii) as well as three sites external to the CCFR: University of Pittsburgh Medical Center (UPMC), Mount Sinai Hospital Toronto, and Seattle-Puget Sound (Access) Cancer Registry [18]. The CCFR enrolled participants after colorectal carcinoma diagnosis with prospective follow-up. The UPMC and Mount Sinai cohorts consisted of consecutively resected colorectal carcinoma at these institutions between 2010 and 2015 and 2011 and 2016, respectively. The Seattle-Puget Sound Cancer Registry cohort has been previously described and consists of patients between 20 and 74 years of age diagnosed with CRC between 2016 and 2018. Recurrence was assessed by manual review of medical records and was available for 3,349 stage I–III <u>CRCs with a median follow-up of 58 months. For the prognostic model training and external validation, the stage</u>

I–III CCFR CRCs with recurrence data (n = 2,411) formed the internal cohort and the UPMC, Mount Sinai, and Seattle-Puget CRCs (n = 938) formed the external validation cohort. This study was approved by the Mayo Clinic institutional review board (IRB 806-96 and 18-11309). No patients were involved in any part of the study, including concept and study design, data collection, analysis and interpretation, drafting of the manuscript, and critical revision. Table 1 shows the characteristics of both train, validation, and test sets that were randomly selected. Figure 1 shows the survival data for both internal and external sites. The p-values are computed using a log-rank test to compare the survival with the internal site Ontario as a reference. Even though Mayo Clinic and UMPC have similar survival rates, we observed that Seattle, Australia, Mt. Sinai, and Access have significantly different survival rates compared to Ontario (based on p-values).

**Table 1.** Description of the dataset - shows the characteristics of both internal and external datasets.

|  |  | Grouped by Data Split |  |  |  |  |
| --- | --- | --- | --- | --- | --- | --- |
|  |  | Overall | external-test | internal-test | internal-train | internal-val |
| n | – | 3411 | 946 | 766 | 1456 | 243 |
| center, n (%) | ACCESS | 128 (3.8) | 128 (13.5) | – | – | – |
|  | Mt. Sinai | 361 (10.6) | 361 (38.2) | – | – | – |
|  | UPMC | 457 (13.4) | 457 (48.3) | – | – | – |
|  | Australia | 233 (6.8) | – | 75 (9.8) | 136 (9.3) | 22 (9.1) |
|  | Hawaii | 37 (1.1) | – | 8 (1.0) | 25 (1.7) | 4 (1.6) |
|  | Mayo | 561 (16.4) | – | 182 (23.8) | 326 (22.4) | 53 (21.8) |
|  | Ontario | 1118 (32.8) | – | 347 (45.3) | 657 (45.1) | 114 (46.9) |
|  | Seattle | 516 (15.1) | – | 154 (20.1) | 312 (21.4) | 50 (20.6) |
| Stage, n (%) | 1 | 689 (20.2) | 209 (22.1) | 151 (19.7) | 278 (19.1) | 51 (21.0) |
|  | 2 | 1364 (40.0) | 376 (39.7) | 292 (38.1) | 601 (41.3) | 95 (39.1) |
|  | 3 | 1358 (39.8) | 361 (38.2) | 323 (42.2) | 577 (39.6) | 97 (39.9) |
| Time to event, mean (SD) |  | 42.2 (20.7) | 38.3 (22.2) | 43.9 (19.9) | 43.8 (19.9) | 42.5 (20.3) |

**Figure 1:**
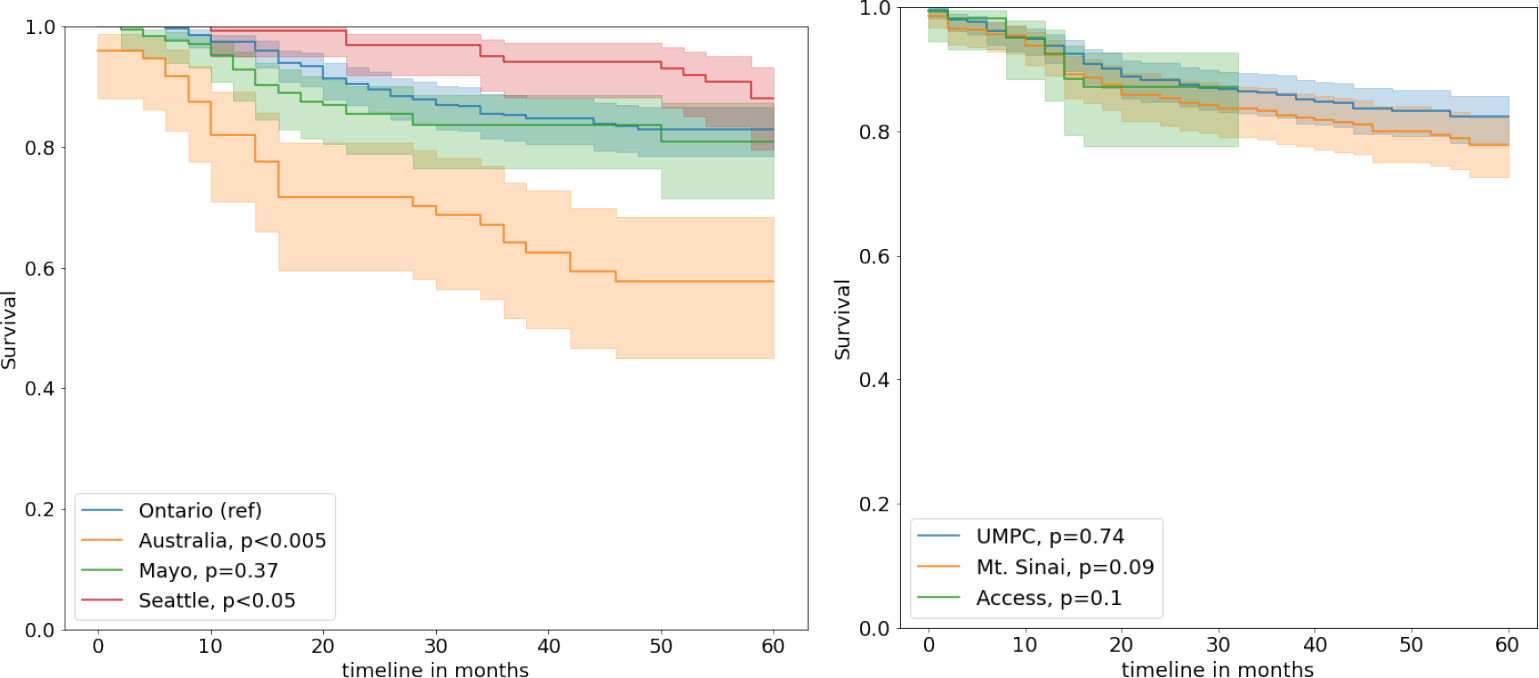
Recurrence free survival for the colorectal carcinoma patients - (a) internal and (b) external sites. P-values are computed as log-rank test using internal site Ontario as reference (ref).

### 2.2. Quantitative image feature extraction

We followed the standard histopathology image acquisition pipeline for image staining and scanning; however, given the geographical distance, the slides were scanned at different locations. From each of the surgically resected colorectal cancer studies, one representative H&E slide was digitized using Leica Aperio GT450 or AT2 at ×40 magnification. The images from the internal cohort were stained with H&E and scanned at Mayo Clinic. The Seattle-Puget cases were stained with H&E at Fred Hutchison Cancer Center and scanned at Mayo Clinic. The UPMC and Mount Sinai cases were stained with H&E and scanned at their respective institutions. After staining, all images were uploaded to the Aiforia Create deep learning cloud-based platform (Aiforia Technologies, Helsinki, Finland), a commercially available platform designed explicitly for histologic images. Each image was manually reviewed by an expert gastrointestinal pathologist (Dr. Pai), and the entire tumor bed was outlined (median 96.5 mm2 analyzed per CRC).

Previously developed deep learning quantitative segmentation algorithm, QuantCRC [19], was applied to the tumor bed to segment colorectal carcinoma digitized images into 13 regions and one object (Figure. 2). The QuantCRC algorithm uses convolutional neural networks (CNN) to segment the image in a stepwise manner and is trained using 24,157 annotations made on 559 images, which are not used in this study. First, the tumor bed is segmented into carcinoma, TB/PDC, stroma, mucin, necrosis, fat, and smooth muscle. The second layer segments stroma into immature (loose, often myxoid stroma with haphazardly arranged plump fibroblasts and collagen fibers), mature (densely collagenous areas with scattered fibroblasts, often with parallel collagen fibers), and inflammatory (dense clusters of chronic inflammatory cells obscuring stromal cells) subtypes. The third layer segments carcinoma into low-grade, high-grade, and signet ring cell carcinoma (SRCC). The fourth layer identifies TILs. To re-validate QuantCRC, 30 images (15 from each GT450 and AT2 scanners) were selected from the 3,349 CRCs with recurrence data. For layers 1 to 3, the algorithm output was compared with annotations by 5 gastrointestinal pathologists. The results from the segmentation algorithm compare favorably with annotations by gastrointestinal pathologists for all features. The most variation was seen between immature and mature stroma, where disagreement among the 5 pathologist raters was noted. This is likely related to the fact that stroma subtyping is a new concept and is not done routinely by pathologists for clinical practice.

**Figure 2:**
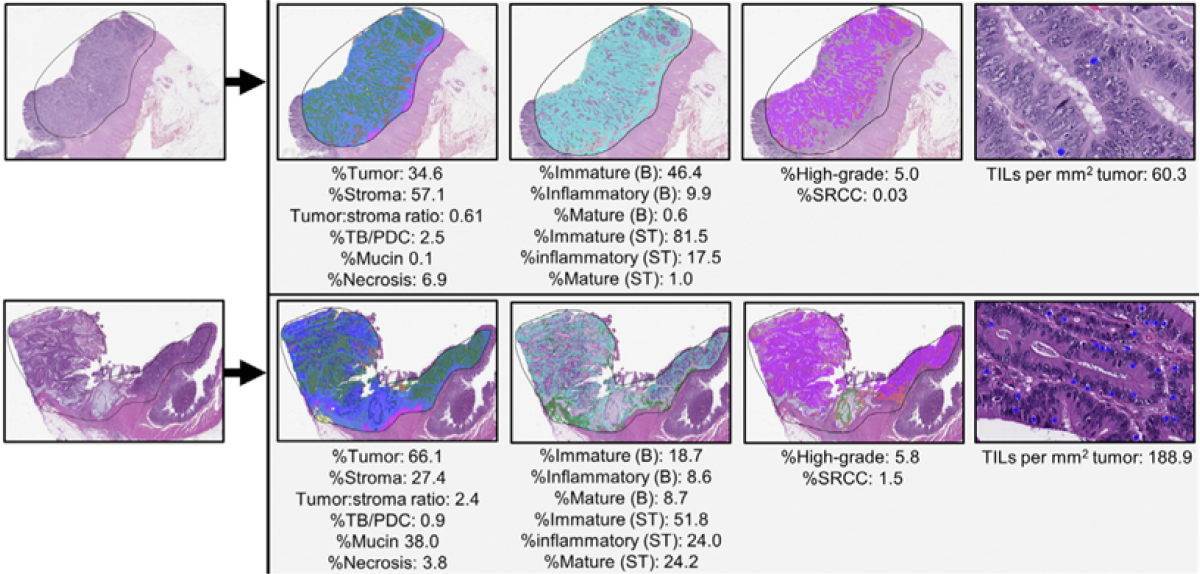
Quantitative feature extraction pipeline on 2 sample images, which segments the image in a stepwise manner. First, the image is segmented into carcinoma (green), stroma (light blue), mucin (dark blue), TB/PDC (red), necrosis (brown), smooth muscle (purple), and fat (yellow). Next, the stroma is segmented into immature (teal), mature (green), and inflammatory (gray). The carcinoma is segmented into low-grade (purple), high-grade (orange), and signet ring cell (light green). Finally, TILs are recognized as objects (blue dots) within the tumor. After this segmentation, 15 features are calculated from each image as shown. Abbreviations: B, tumor bed; ST, stromal region.

For each image, fat and smooth muscle were subtracted from the tissue area to generate the tumor bed area. From the tumor bed area, the following 15 quantitative parameters were measured: %tumor, %stroma, tumor:stroma ratio, %TB/PDC within the tumor, %mucin within the tumor, %necrosis within the tumor bed, %high-grade, %SRCC, TILs per mm2 of tumor, %immature stroma (tumor bed), %inflammatory stroma (tumor bed), % mature stroma (tumor bed), %immature (stromal region), % inflammatory (stromal region), and %mature (stromal region). These 15 quantitative parameters comprise the QuantCRC features used for downstream analysis.

### 2.3. Causal survival model

Figure 3 presents the proposed causal modeling framework for recurrence free survival estimation across multiple sites. Our primary hypothesis is that by using the proxy (*W*) and auxiliary (*C*) variable in a causal framework, we Figure 3: Causal modeling framework: (left) latent shift assumption and causal relations; (right) proposed model diagram demonstrates 3 main components of the model. A) learns a latent representation capturing task relevant information alongside proxy information. B) Attempts to infer the latent variables directly from the input features. C) A risk estimation model is trained, predictions are modified using the latent estimates will be able to adapt to the latent subgroup shift that appears between sites without knowing the cause of bias and able to predict the survival *Y* given the input *X* (Figure 3.i). Following [17], if we have two domains (*P* source and *Q* target), we frame the learning *Q*(*Y* |*X*) as identification problem where observations drawn from *P* (*X, C, Y, W*) and *Q*(*X*). We assume that *C* and *W* are observed in the source distribution and thus can be used during learning when *U* is unobserved and *U* is a discrete variable. We also assume that conditional dependencies in the data exist iff they exist in the graph (Figure 3.i). Note that we only observe (*X, C, Y, W*) in the source *P* and *X* in the target *Q*.

**Figure 3:**
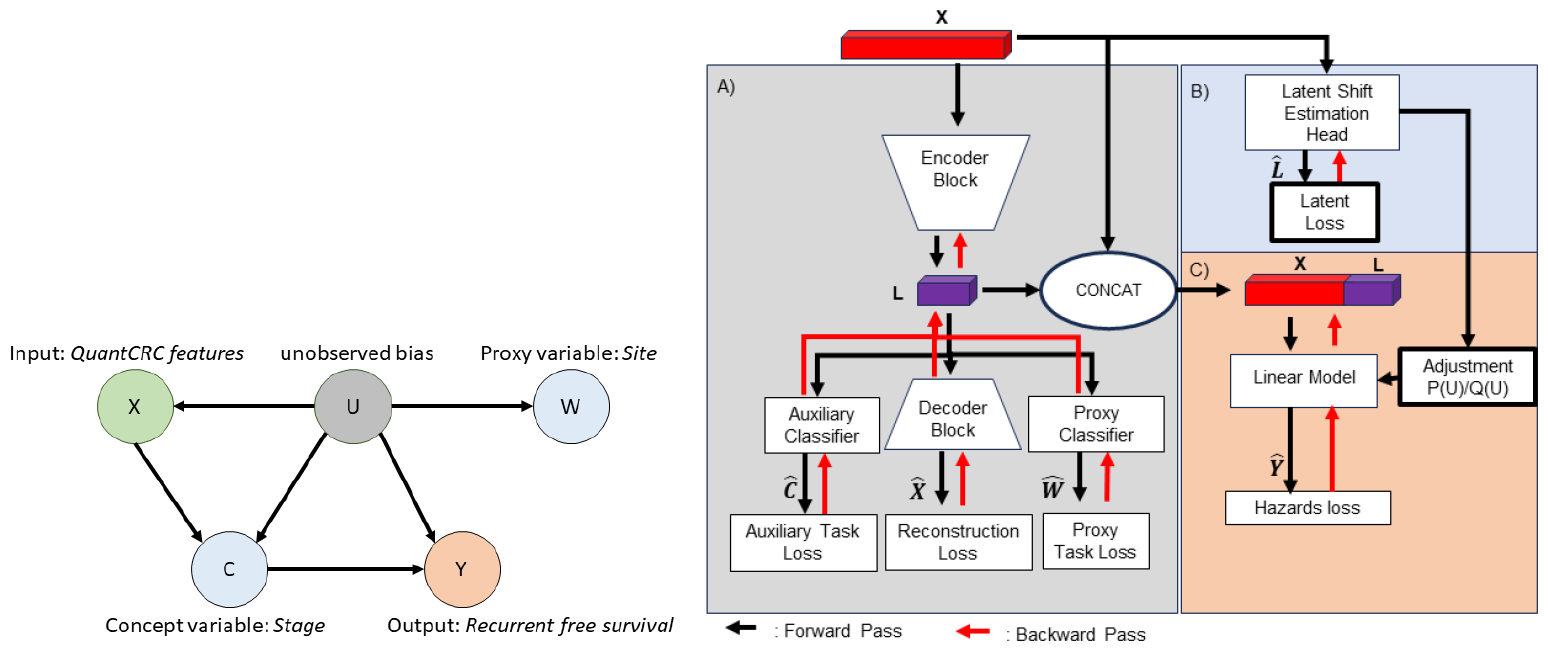
Causal modeling framework: (left) latent shift assumption and causal relations; (right) proposed model diagram demonstrates 3 main components of the model. A) learns a latent representation capturing task relevant information alongside proxy information. B) Attempts to infer the latent variables directly from the input features. C) A risk estimation model is trained, predictions are modified using the latent estimates

We adopted the latent shift causal framework for predicting recurrence-free survival (*Y*) of colorectal carcinoma patients using the quantitative histopathology features (*X*). Given the issue of not knowing the unobserved bias variable, we model the site of treatment as a proxy (*W*) and the cancer stage as a concept (*C*). We assume that input histopathology features (*X*) are affected by the site due to the bias in acquisition and population variations, and cancer stage directly effects recurrence-free survival (*Y*). The learning is being done using three core components (Figure 3).

First, in module A, we learn the unobserved bias variable *U* by training an autoencoder to reconstruct the input, i.e. QuantCRC features, and predict the proxy and concept. Thus, in addition to the reconstruction loss for the features, the encoder component is also trained with two auxiliary classifiers - (i) classify tumor staging (classify concept vector *C*), and (ii) predict hospital of origin (predict the proxy *W*). The classification task uses categorical cross-entropy losses. The learning of the autoencoder model is constrained to predict a discrete latent space to ultimately represent the latent variable *U*. We incorporated Wasserstein auto encoder to reduce prior-collapse.

During the second stage, in module B, the auto-encoder component is frozen, and a parallel latent estimation branch is trained to predict latent variables *U* from the original QuantCRC features directly without referring the proxy and concept loss. The estimation of the latent is used during the final adaptation phase to estimate the latent shift and adjust the model’s final risk predictions.

In the final stage of the training process (module C), we train a final risk prediction model that leverages the QuantCRC features (*X*) and learned latent variable *U* (module B) to predict the risk of recurrence at different time points. Using the QuantCRC features of the external data (domain Q), the latent estimation branch is used to predict latent variables, and a shift ratio is then estimated. The estimates from the training data (domain *P*) and external data (domain *Q*) are compared, and the difference is used to re-scale the model predictions (see equation 1). The recurrence risk estimation head (module C) is then fine-tuned using the internal data (*P*) to account for the new latent shift estimations based on an estimate of *U*.

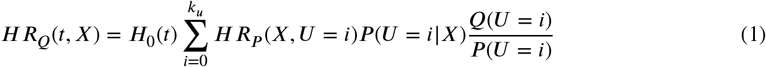

During inference, the model use the predicted latent shift of the new data to re-scale predictions. For our use case, we will treat samples from the external dataset (*Q*) as a single domain for which we adjust our model predictions. All models were trained using the same training parameters: batch size of 256, Adam optimizer with a learning rate of 0.01, and early stopping based on validation loss.

## 3. Results

Figure 4 shows the pair-wise Pearson correlation coefficient values between QuantCRC features, stage, center, and recurrence. As we can observe that individual features do not have high correlation (*>* 0.5) with recurrence (*Y*); however, stroma and tumor bed-related features are highly correlated among themselves and also moderately correlated with center and stage. Interestingly, combination of center and stage increase the correlation with the target recurrence *Y*, which follows our hypothesis that the unobserved variable (*U*) that effects the outcome (*Y*) can be estimated from stage (*C*) and center (*W*).

**Figure 4:**
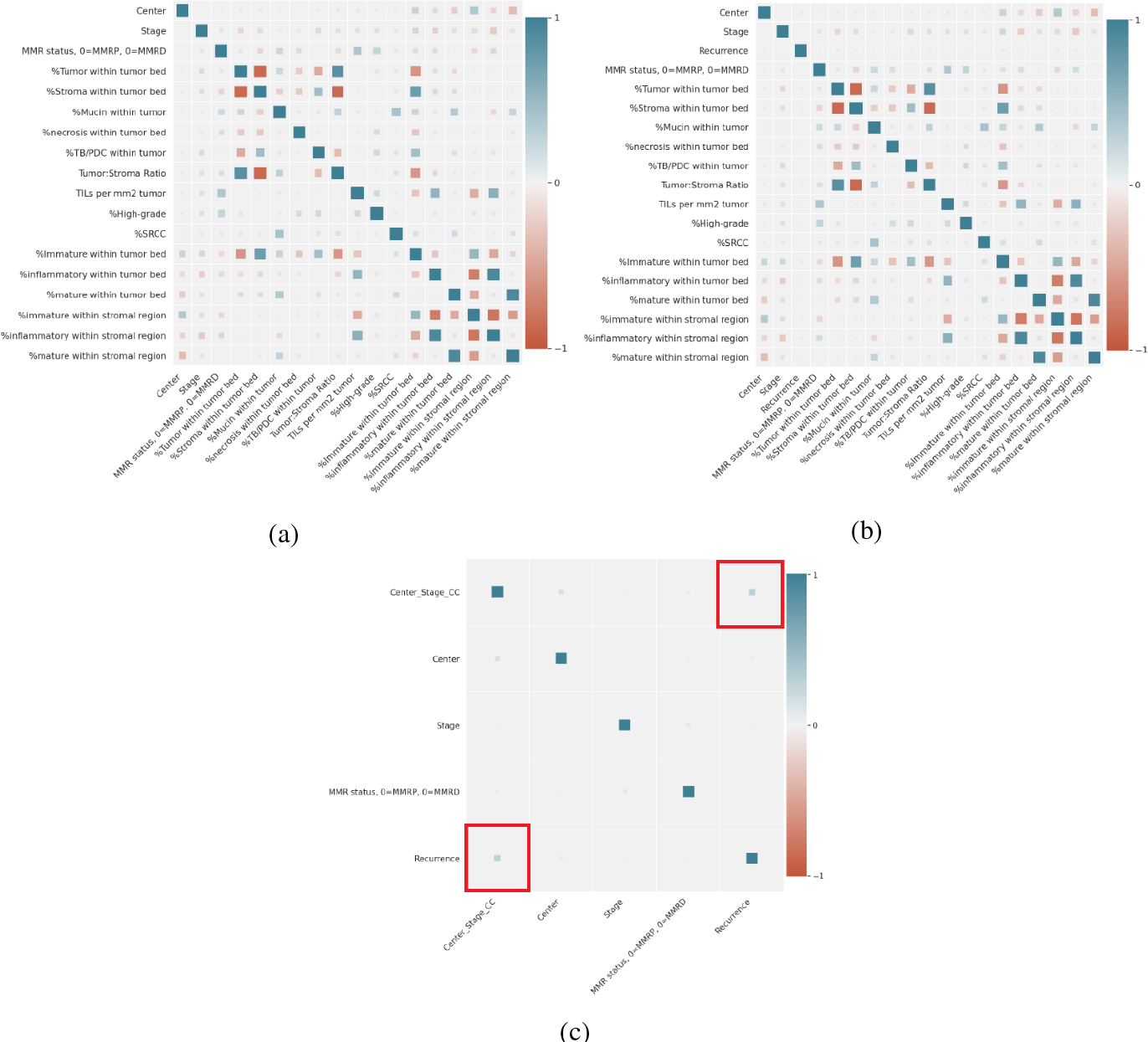
Pearson correlation coefficient between Stage (*C*), Center (*W*), QuantCRC features (*X*) and target recurrence (*Y*) - (a) represents stage and center as separate variables, (b) represents stage and center as a combined variables, and (c) Correlation with recurrence.

As baseline, we compared our proposed model performance against traditional Cox Proportional Hazard (CPH) model trained using the lifelines package [20] and DeepSurv model [21] - a multi-layer deep neural network with hazard loss. Model evaluation was done by measuring the concordance index of the model’s risk score at 60 months, comparing the ranking of at-risk patients adequately. The 95% confidence interval was obtained for all the model performances using the autobootstraping. Overall and site-based performance is presented in Table 2.

**Table 2.**
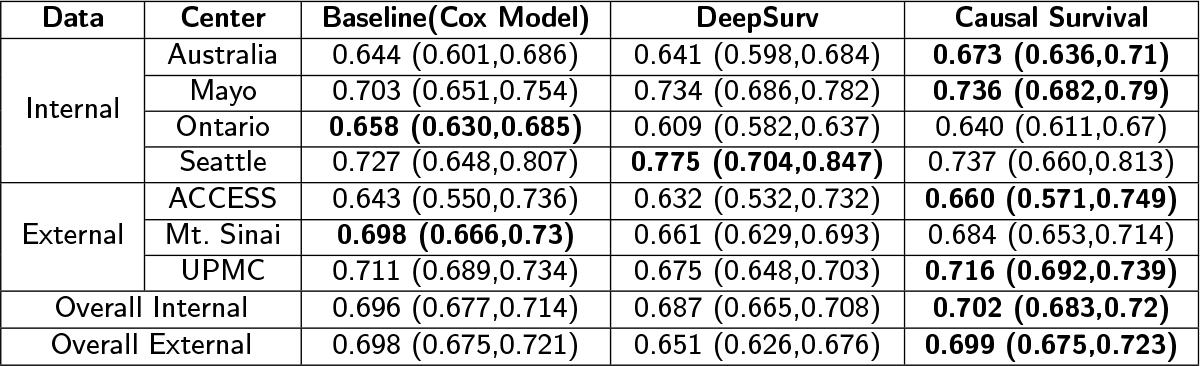
Performance of the models on the internal and external test sets in-terms of C-index. 95% confidence interval is calculated using auto-bootstraping. **Bold** front represents optimal performance.

Overall, the CPH baseline (CI internal: 0.696, external:0.698) and Causal Survival (CI internal:0.702, external:0.699) models achieve comparable performance on the overall internal and external test sets while DeepSurv performance is consistently lower with higher variability. However, the performance differences between the sites are significant in the baseline Cox model, e.g., C-index for Seattle is 0.727, and for Australia is 0.644. While maintaining the overall performance, the Causal survival model reduces the disparities between the site performance, e.g., the C-index for Seattle is 0.737, and for Australia is 0.673. Similar results can be observed for the external dataset that the Causal survival model improved the performance for all the sites, including ACCESS.

In Figure 5, we represent comparative performance between the models in terms of the area under the operating characteristics curve (AUROC) at different time intervals. Causal survival model performance is consistently better for all the time points on the internal dataset, except 2 months, which has the lowest rate of recurrence. A similar consistent performance boost was observed for the external data for predicting short-term as well as long-term recurrence. On the external data, C ausal survival model has a huge performance boost over DeepSurv (≈ 5% AUC).

**Figure 5:**
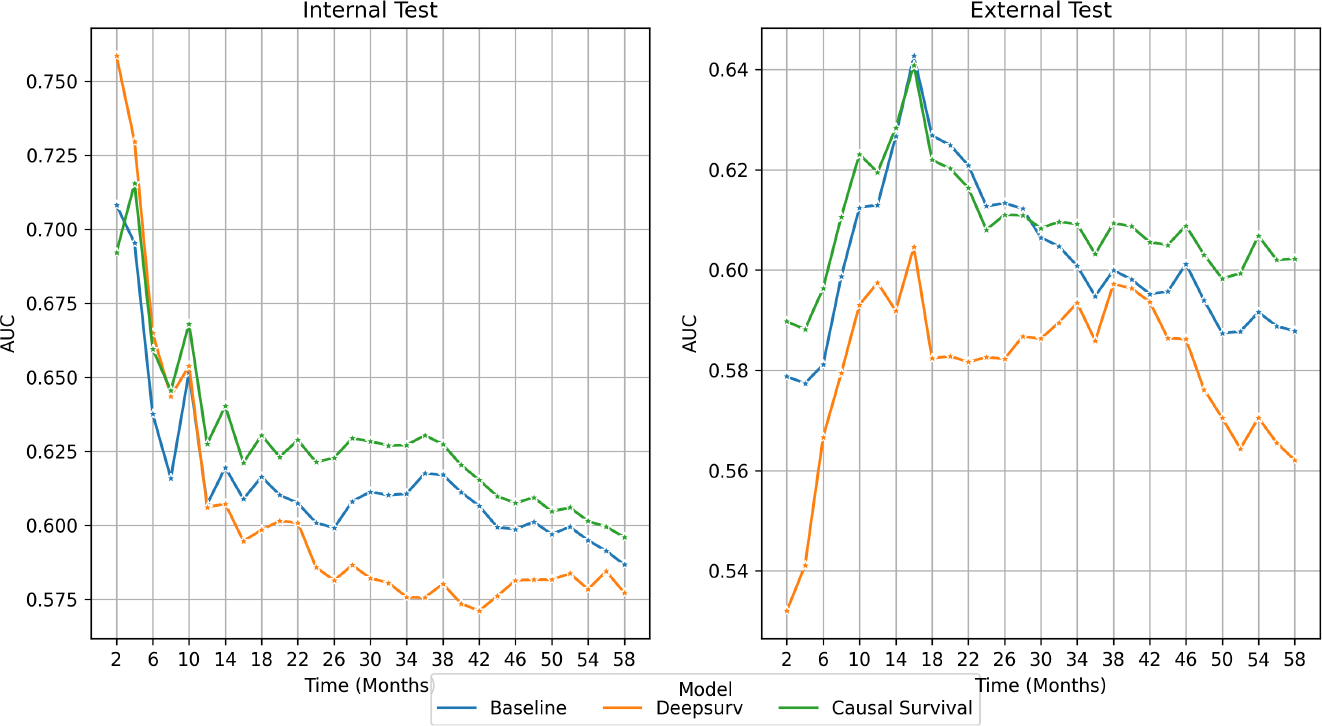
Comparative Area under the receiver operating characteristics curve (AUROC) calculated for every time step on the internal and external dataset - (left) internal and (right) external. Blue: Baseline Cox model, Orange: DeepSurv, and Green: Proposed Causal Survival model.

### 3.1. Ablation

To demonstrate the efficacy of our proposed adaptation of the causal domain adaptation framework, the modularized framework allows us to evaluate the effect of key modules on the final predictions. Table 3 highlights the abalation performance for different configurations.

**Table 3.** Configuration of ablation study. Removed components of the causal survival model is marked as X. Performance measured as C-index with 95% confidence interval.

| Latent Shift Probability Adjustment | Infer Using Latent Variable | Infer using Quant CRC Features | Internal-Test Concordance Index | External Test Concordance Index |
| --- | --- | --- | --- | --- |
| X |  |  | 0.712 (0.694,0.73) | 0.691 (0.669,0.713) |
|  | X |  | 0.538 (0.513,0.562) | 0.486 (0.462,0.51) |
|  |  | X | 0.702 (0.683,0.721) | 0.686 (0.661,0.711) |

*i.Removal of Latent Shift Adjustment (dropped module C)*:

First, we remove the proposed domain adjustment component from module C, i.e. *P* (*U*)/*Q*(*U*) and evaluate the performance on the internal and external test data. As expected, removal of latent shift domain adjustment does actually improved the performance on the internal data (*P*) but dropped the performance on the external test (*Q*). This shows the fact that proposed latent shift adjustment helps to improve the generalizability of the model on the external data.

*ii. Infer only using Latent Variable (dropped module B and C)*:

As the second experiment, we only use the latent variable derived by module A and pass it through hazard layer to compute the survival risk. The performance of the model remains high in the internal test which demonstrates the latent features from the encoder are able to capture task relevant correlation. Given the latent estimation head and the adjustment parameter for the domain *P* is missing the performance dropped on the external datasets.

*iii. Infer only using QuantCRC fetaures (dropped module A, B and C)*:

The final experiment we only used the QuantCRC features. We observed that the model performance decrease to be in range with the Deepsurv model in the internal test set. Performance on the external set Drops to the lowest amongst all models. Suggesting the predictive power of the latent features adjusted based on proxy and concept is essential for proper risk prediction.

## 4. Discussion

Bias in AI models for histopathology domain can arise from several sources - (i) *Data bias*: If the training data is not representative of the whole population, the model may not generalize well on new data. For example, if the training data only includes samples from one scanner, the model may not perform well on samples from other acquisition device; (ii) *Algorithmic bias*: The algorithms used to train the model may introduce bias. For example, if the algorithm is designed to optimize for a specific metric (e.g. AUC), it may ignore other important factors. such as disparities between the ethnic subgroups; (iii) *Human bias*: Human bias can be introduced during the data annotation process or when selecting the data used to train the model. It is important to identify and address these sources of bias to ensure that the AI model performs accurately and fairly. However, often cause of the bias stays unknown as it can be simultaneously affected by multiple factors.

We proposed a Causal Survival model that can reduce the effect of unknown bias via a causal reasoning framework incorporated within a deep learning paradigm. As our first use case, we evaluated the model for predicting recurrence for colorectal cancer patients across seven geographically distributed sites using the Colon Cancer Family Registry (CCFR) and showed improvement in performance across sites. Our primary contribution is to adapt the unsupervised domain adaptation technique to adjust the deep latent space of the out-of-domain samples and ultimately obtain analogus performance across in-domain and out-of-domain samples.

Interestingly, as shown in the correlation plots (Figure 4), cancer stage and recurrence-free survival across sites has limited to no correlation. Indicating that other factors beyond staging can mediate the risk of recurrence for the colorectal cancer patients. Our causal modeling approach suggests that the models are able to learn measure of the hidden variations in data, in other words ‘unknown bias’, and utilize it to improve prediction generalizability and efficiently handle distribution shifts between internal and external data. In particular, we observe that survival rate from Australia, Seattle, Mt. Sinai and Access are significantly different (*p <* 0.1) from the reference internal site Ontario. Additionally, there is a positive correlation between linear combination of center and stage, with the recurrence free survival. The disparity is reflected in the models’ performance where the baseline models (Cox and DeepSurv) underperformed or overperformed on these sets. While our proposed architecture demonstrates an overall improvement on both internal and external datasets. We observe a significant improvement in the risk prediction on the three external datasets without utilizing fine-tuning or data harmonization techniques. Our ablation study demonstrated that by learning the latent features with auxiliary losses defined based on measured potential bias factors, an improvement in model predictions can be obtained. Finally, incorporating the latent shift adjustment further guarantees the stability of model predictions in a new domain.

The proposed framework is an adaption of unsupervised latent distribution shift method where we introduce a deep latent space features and a domain adjustment factor. We also extend the framework for prognosis of recurrence-free survival for Colorectal Cancer patients from quantitative histopathology features. Though detection and interpretation of ‘unknown bias’ is still an unsolved challenge, our proposed solution estimate the bias via the proxy and concept variable and reduce the bias to improve AI model generalizibility.

## Limitation

The study has several limitations. First, the Causal Survival framework requires the availability of two mediating concepts *C* and of a proxy variable *W* at training time. This measures might not be readily available for other studies, or it may not satisfy all the assumptions described in the framework. Furthermore, the causal assumptions are typically not testable as *U* is not observed. Second, we validated the model on the quantified histopathology features but ideally the paradigm can also be used on the raw image data. Third, we only observed a moderate improvement of performance which could be due to fact that the slides are scanned and digitized using a standard protocol to reduce variations between the scanned slides within CCFR. Furthermore, the manual annotation variations are reduced by a single expert gastrointestinal pathologist drawing the tumor boundary. We believe that the site specific variations and the bias will be amplified in a scenario where multiple sites are contributing the clinically scanned slides independently without a common protocol. In such cases, performance improvement over the baseline survival models will be more beneficial with the proposed method.

## Data Availability

Data cannot be shared publicly because of the data privacy requirement. Data are available from the CCFR for researchers who meet the criteria for access to confidential data.

## References

[1] K. Ashman, H. Zhuge, E. Shanley, S. Fox, S. Halat, A. Sholl, B. Summa, J. Q. Brown, Whole slide image data utilization informed by digital diagnosis patterns, Journal of Pathology Informatics 13 (2022) 100113.

[2] A. Madabhushi, G. Lee, Image analysis and machine learning in digital pathology: Challenges and opportunities, Medical image analysis 33 (2016) 170–175.

[3] D. Bousis, G.-I. Verras, K. Bouchagier, A. Antzoulas, I. Panagiotopoulos, A. Katinioti, D. Kehagias, C. Kaplanis, K. Kotis, C.-N. Anagnostopoulos, et al., The role of deep learning in diagnosing colorectal cancer, Gastroenterology Review/Przegląd Gastroenterologiczny 18 (2023).

[4] C. Zhang, S. Bengio, M. Hardt, B. Recht, O. Vinyals, Understanding deep learning (still) requires rethinking generalization, Communications of the ACM 64 (2021) 107–115.

[5] I. Banerjee, K. Bhattacharjee, J. L. Burns, H. Trivedi, S. Purkayastha, L. Seyyed-Kalantari, B. N. Patel, R. Shiradkar, J. Gichoya, “shortcuts” causing bias in radiology artificial intelligence: causes, evaluation and mitigation., Journal of the American College of Radiology (2023).

[6] L. Seyyed-Kalantari, H. Zhang, M. B. McDermott, I. Y. Chen, M. Ghassemi, Underdiagnosis bias of artificial intelligence algorithms applied to chest radiographs in under-served patient populations, Nature medicine 27 (2021) 2176–2182.

[7] F. M. Howard, J. Dolezal, S. Kochanny, J. Schulte, H. Chen, L. Heij, D. Huo, R. Nanda, O. I. Olopade, J. N. Kather, et al., The impact of site-specific digital histology signatures on deep learning model accuracy and bias, Nature communications 12 (2021) 4423.

[8] D. Komura, S. Ishikawa, Machine learning methods for histopathological image analysis, Computational and structural biotechnology journal 16 (2018) 34–42.

[9] E. Reinhard, M. Adhikhmin, B. Gooch, P. Shirley, Color transfer between images, IEEE Computer graphics and applications 21 (2001) 34–41.

[10] M. Macenko, M. Niethammer, J. S. Marron, D. Borland, J. T. Woosley, X. Guan, C. Schmitt, N. E. Thomas, A method for normalizing histology slides for quantitative analysis, 2009 IEEE international symposium on biomedical imaging: from nano to macro (2009) 1107–1110.

[11] D. Tellez, M. Balkenhol, I. Otte-Höller, R. van de Loo, R. Vogels, P. Bult, C. Wauters, W. Vreuls, S. Mol, N. Karssemeijer, et al., Whole-slide mitosis detection in h&e breast histology using phh3 as a reference to train distilled stain-invariant convolutional networks, IEEE transactions on medical imaging 37 (2018) 2126–2136.

[12] J. R. Zech, M. A. Badgeley, M. Liu, A. B. Costa, J. J. Titano, E. K. Oermann, Variable generalization performance of a deep learning model to detect pneumonia in chest radiographs: a cross-sectional study, PLoS medicine 15 (2018) e1002683.

[13] J. Rueckel, L. Trappmann, B. Schachtner, P. Wesp, B. F. Hoppe, N. Fink, J. Ricke, J. Dinkel, M. Ingrisch, B. O. Sabel, Impact of confounding thoracic tubes and pleural dehiscence extent on artificial intelligence pneumothorax detection in chest radiographs, Investigative Radiology 55 (2020) 792–798.

[14] J. W. Gichoya, I. Banerjee, A. R. Bhimireddy, J. L. Burns, L. A. Celi, L.-C. Chen, R. Correa, N. Dullerud, M. Ghassemi, S.-C. Huang, et al., Ai recognition of patient race in medical imaging: a modelling study, The Lancet Digital Health 4 (2022) e406–e414.

[15] L. Seyyed-Kalantari, G. Liu, M. McDermott, I. Y. Chen, M. Ghassemi, Chexclusion: Fairness gaps in deep chest x-ray classifiers, in: BIOCOMPUTING 2021: proceedings of the Pacific symposium, World Scientific, 2020, pp. 232–243.

[16] R. Correa, M. Shaan, H. Trivedi, B. Patel, L. A. G. Celi, J. W. Gichoya, I. Banerjee, A systematic review of ‘fair’ai model development for image classification and prediction, Journal of Medical and Biological Engineering 42 (2022) 816–827.

[17] I. Alabdulmohsin, N. Chiou, A. D’Amour, A. Gretton, S. Koyejo, M. J. Kusner, S. R. Pfohl, O. Salaudeen, J. Schrouff, K. Tsai, Adapting to latent subgroup shifts via concepts and proxies, in: International Conference on Artificial Intelligence and Statistics, PMLR, 2023, pp. 9637–9661.

[18] P. A. Newcomb, J. Baron, M. Cotterchio, S. Gallinger, J. Grove, R. Haile, D. Hall, J. L. Hopper, J. Jass, L. Le Marchand, et al., Colon cancer family registry: an international resource for studies of the genetic epidemiology of colon cancer, Cancer Epidemiology Biomarkers & Prevention 16 (2007) 2331–2343.

[19] R. K. Pai, D. Hartman, D. F. Schaeffer, C. Rosty, S. Shivji, R. Kirsch, R. K. Pai, Development and initial validation of a deep learning algorithm to quantify histological features in colorectal carcinoma including tumour budding/poorly differentiated clusters, Histopathology 79 (2021) 391–405.

[20] C. Davidson-Pilon, lifelines: survival analysis in python, Journal of Open Source Software 4 (2019) 1317.

[21] J. L. Katzman, U. Shaham, A. Cloninger, J. Bates, T. Jiang, Y. Kluger, Deepsurv: personalized treatment recommender system using a cox proportional hazards deep neural network, BMC medical research methodology 18 (2018) 1–12.

